# Antibiotic Consumption Patterns among Outpatients at a Cardiac Tertiary Hospital in Tanzania

**DOI:** 10.1101/2025.11.18.25340496

**Authors:** Hamis Abdalla Kaniki, Raphael Z. Sangeda, Jacob Aron Ndayomwami, James Mwakyomo, Naizihijwa Majani, Reuben Kato Mutagaywa, Adelard Bartholomew Mtenga, Peter Richard Kisenge

## Abstract

**Background:** Outpatient encounters are significant contributors to avoidable antibiotic exposure. However, the dispensing-based outpatient consumption data for Tanzania are limited. Most national estimates rely on import or sales data that do not reflect patient-level use. Cardiovascular clinics are particularly relevant because cardiopulmonary symptom overlap can lead to precautionary prescribing.

**Methods:** We conducted a retrospective dispensing-based utilization analysis of cardiovascular outpatients at the Jakaya Kikwete Cardiac Institute (JKCI) in Dar es Salaam, Tanzania, from July 2017 to June 2022. Systemic antibacterial agents were mapped to the World Health Organization anatomical therapeutic chemical/defined daily dose (WHO ATC/DDD) and expressed as defined daily doses per 100 outpatients per day. Consumption patterns were summarized by fiscal year, molecule, ATC level 3, and WHO Access/Watch/Reserve (AWaRe) category. Monthly consumption was decomposed to evaluate trends and seasonality.

**Results:** A total of 11,586 outpatients received at least one systemic antibiotic during the study period. Annual outpatient antibiotic utilization remained low and stable, ranging from approximately 0.27 to 0.38 DDD per 100 outpatients per day across fiscal years, with a cumulative total of 1.68 DDD per 100 outpatients per day across the five years. Consumption was dominated by beta-lactam/penicillin antibiotics, with Access antibiotics accounting for 48.5% of cumulative exposure, followed by Watch antibiotics (45.5%) and minimal Reserve use (5.9%). However, the Access proportion was below the WHO Access-60 benchmark for optimal outpatient use. Time-series decomposition revealed a modest upward trend, peaking in early 2021, accompanied by a recurring seasonal pattern of monthly use.

**Conclusion:** Outpatient antibiotic utilization at JKCI was quantitatively low, yet showed persistent exposure dominated by Access antibiotics. Dispensing-based outpatient surveillance is feasible and should be integrated into routine stewardship dashboards to target avoidable outpatient antibiotic use.

## Introduction

Antibiotics are essential for preventing and treating bacterial infections; however, inappropriate use accelerates antimicrobial resistance (AMR), leading to increased morbidity, mortality, and healthcare costs [1]. Surveillance of antibiotic consumption is a core component of global AMR strategies. In Tanzania, the National Action Plan on AMR prioritizes monitoring antimicrobial utilization and promoting rational prescribing at all levels of care [2–5]. Despite these priorities, operational data on outpatient antibiotic consumption in Tanzania remain sparse. Most national estimates rely on importation and sales datasets [5–10], which reflect market volumes rather than antibiotics dispensed to individual patients. While some studies have described longitudinal inpatient antibiotic use using electronic medical records, point prevalence surveys are only recently being conducted in other low- and middle-income countries (LMICs) [4,12–23].

Cardiology outpatient encounters are common, and symptoms such as cough, throat irritation, chest discomfort, and dyspepsia can overlap with cardiac presentations, increasing the risk of precautionary or unnecessary antibiotic use. International evidence indicates that outpatient settings contribute significantly to community antibiotic exposure and often serve as major reservoirs of avoidable use [24,25]. Similar outpatient studies in LMICs reported substantial ambulatory antibiotic exposure, even when bacterial infection was unlikely [26–29]. However, the Access share of 48.5% remained below the WHO Access-60 benchmark, indicating the scope for further improvement in outpatient prescribing optimization. Outpatient consumption analyses from Syria and Ethiopia have shown high utilization of broad-spectrum antibiotics at the primary care level [26,28]. At the same time, findings from Ethiopia and Nigeria have demonstrated that stewardship interventions can substantially reduce unnecessary prescriptions [27,29]. Specialist facility trend analyses also indicated measurable correlations between outpatient antibiotic consumption and AMR trajectories over time [30]. These patterns suggest that outpatient consumption, even in non-infectious clinical specialties, remains a meaningful target for stewardship investment.

However, no study in Tanzania has quantified outpatient antibiotic utilization using the dispensing-based World Health Organization Anatomical Therapeutic Chemical/Defined Daily Dose (WHO ATC/DDD) methodology in a clinical hospital setting.

Unlike national antimicrobial consumption metrics, which typically use DDD per 1,000 inhabitants per day (DID), patient-level dispensing in outpatient clinics requires a patient-specific denominator. Therefore, in this analysis, antibiotic consumption was expressed as DDD per 100 outpatients per day, thereby normalizing utilization to the number of outpatients actually seen at the facility.

This study quantified systemic antibiotic consumption among cardiovascular outpatients at the Jakaya Kikwete Cardiac Institute (JKCI) in Dar es Salaam using the WHO ATC/DDD methodology, characterized temporal patterns including WHO Access, Watch, and Reserve (AWaRe) class distributions, and examined variation in consumption across demographic subgroups to identify priority targets for outpatient antimicrobial stewardship.

## Materials and methods Study design and setting

We conducted a retrospective dispensing-based utilization study at the Jakaya Kikwete Cardiac Institute (JKCI) in Dar es Salaam, Tanzania. JKCI is a national public cardiovascular referral center co-located within Muhimbili National Hospital, with approximately 103 inpatient beds and an average outpatient volume of around 700 patients per week. The study period spanned from July 2017 to June 2022.

## Population and data source

Dispensing records for all medications were extracted from the JKCI MedPro pharmacy database. The variables available included patient identifier, sex, age, date of dispensing, visit identifier, inpatient/outpatient status, mode of payment, pharmaceutical item description, quantity dispensed, dosage form, and pharmacy outlet. "Outpatients" included all encounters without an assigned bed. This analysis was restricted to outpatients who were dispensed with systemic antibacterials (oral or parenteral). Patients taking topical antibiotics, antivirals, antifungals, antiparasitics, or anti-tuberculosis agents were excluded.

## ATC/DDD mapping

Each antibiotic record was harmonized to generic name, route, strength, and unit, mapped to WHO ATC codes (levels 5 to 1), and assigned a DDD value using the WHO Collaborating Center for Drug Statistics Methodology. The total grams dispensed were converted to DDDs.

## Consumption metric

Outpatient facility-level analysis utilization was expressed as DDD per 100 outpatients per day. This denominator is anchored to the actual number of cardiovascular patients seen at the JKCI. Therefore, it was calculated using the following formula:

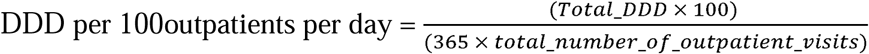

For interpretive clarity, the DDD values presented in the main text were rounded to two decimal places, while the underlying calculations retained full precision.

## Classifications

Utilization was summarized by route, dosage form, ATC classes, individual molecules, and WHO Access/Watch/Reserve (AWaRe) categories. Additionally, the Drug Utilization 50 and 90 (DU50/DU90) indicators were computed by ranking level 5 ATC molecules by cumulative annual DDD contribution and determining the number of molecules that cumulatively account for 50% and 90% of total outpatient antibiotic consumption.

## Statistical analysis

Data cleaning and aggregation were performed in Microsoft Power BI. Group comparisons were conducted using the χ² test. Time-series decomposition of monthly utilization (trend, seasonality, and residuals) was conducted using IBM SPSS version 26 with an additive decomposition model. Statistical significance was set at *p*<0.05.

## Ethics

Ethical approval was obtained from the Muhimbili University of Health and Allied Sciences Institutional Review Board (Ref: DA.25/111/01; approval date: February 8, 2021). Only de-identified data were used, and confidentiality was maintained.

## Results

A total of 11,586 cardiovascular outpatients were included in this analysis from the Jakaya Kikwete Cardiac Institute (JKCI) in Dar es Salaam, Tanzania, representing a cumulative 1.68 DDD per 100 outpatients per day over the five years. Females accounted for 59.5% (N=6,897), while males accounted for 40.5% (N=4,689). Older adults were the dominant group, accounting for 49.3% (N=5,716) of participants aged ≥50 years. NHIF coverage was the primary financing mechanism, accounting for 72.4% (N = 8,385). Most patients received antibiotics within a single fiscal year, with 65.5% (N = 7,588) (Table 1).

**Table 1:** Antibiotic consumption among cardiovascular outpatients at the Jakaya Kikwete Cardiac Institute (JKCI), Dar es Salaam, Tanzania (N = 11,586)

| Variable | Category | N (%) | Mean DDD per 100 outpatients/day (± SD) | Sum DDD/100 | p-value |
| --- | --- | --- | --- | --- | --- |
| **Sponsor Group** | AAR | 41 (0.4%) | 0.0001418 ($\pm 0.0002036$) | 0.01 | <0.001 |
| | Private | 2710 (23.4%) | 0.0000832 ($\pm 0.0001892$) | 0.23 | |
| | Exemption | 141 (1.2%) | 0.0001173 ($\pm 0.0001298$) | 0.02 | |
| | Military | 67 (0.6%) | 0.0001692 ($\pm 0.0001351$) | 0.01 | |
| | Jubilee Insurance | 25 (0.2%) | 0.0002387 ($\pm 0.0003478$) | 0.01 | |
| | NHIF | 8385 (72.4%) | 0.0001667 ($\pm 0.0002737$) | **1.40** | |
| | NSSF | 131 (1.1%) | 0.0001704 ($\pm 0.0001864$) | 0.02 | |
| | SIC Insurance | 54 (0.5%) | 0.0001939 ($\pm 0.0003423$) | 0.01 | |
| | Zanzibar (SMZ) | 18 (0.2%) | 0.0001047 ($\pm 0.0001608$) | 0.00 | |
| | Other | 7 (0.1%) | 0.0000569 ($\pm 0.0000633$) | 0.00 | |

| Variable | Category | N (%) | Mean DDD per 100<br>outpatients/day (± SD) | Sum<br>DDD/100 | p-value |
| --- | --- | --- | --- | --- | --- |
| **Sex** | Male | 4689<br>(40.5%) | 0.0001311 (±0.0002375) | 0.61 | <0.001 |
| | Female | 6897<br>(59.5%) | 0.0001572 (±0.0002673) | 1.08 | |
| **Age group** | 0–9 | 2754<br>(23.8%) | 0.0000879 (±0.0002569) | 0.24 | <0.001 |
| | 10–19 | 646<br>(5.6%) | 0.0000951 (±0.0001399) | 0.06 | |
| | 20–29 | 564<br>(4.9%) | 0.0001756 (±0.0002631) | 0.10 | |
| | 30–39 | 818<br>(7.1%) | 0.0002260 (±0.0003466) | 0.18 | |
| | 40–49 | 1088<br>(9.4%) | 0.0001946 (±0.0003461) | 0.21 | |
| | 50–59 | 1752<br>(15.1%) | 0.0001734 (±0.0002711) | 0.30 | |
| | 60–69 | 2075<br>(17.9%) | 0.0001521 (±0.0001791) | 0.32 | |
| | 70 and above | 1889<br>(16.3%) | 0.0001485 (±0.0002081) | 0.28 | |
| **Fiscal Year** | 2017–2018 | 2225<br>(19.2%) | 0.0002519 (±0.0004131) | 0.56 | <0.001 |
| | 2018–2019 | 2201<br>(19.0%) | 0.0001441 (±0.0002213) | 0.32 | |
| | 2019–2020 | 2307<br>(19.9%) | 0.0001466 (±0.0002502) | 0.34 | |
| | 2020–2021 | 2353<br>(20.3%) | 0.0001163 (±0.0001561) | 0.27 | |
| | 2021–2022 | 2500<br>(21.6%) | 0.0000838 (±0.0001159) | 0.21 | |
| **Lost first**<br>**year** | No: continued into the<br>following years | 3998<br>(34.5%) | 0.00023766 (±0.00035377) | 0.95 | <0.001 |
| | Yes: started & ended<br>same year <2022 | 7588<br>(65.5%) | 0.00009867 (±0.00016574) | 0.75 | |

Antibiotic utilization differed significantly across patient subgroups (all p < 0.001). NHIF accounted for 1.40 DDD per 100 outpatients per day (80.0% DDD), while private payers accounted for 0.23 DDD per 100 outpatients per day (13.1% DDD). Females contributed 1.08 DDD per 100 outpatients per day (63.9% DDD) compared with 0.61 DDD per 100 outpatients per day (36.1% DDD) among males. Earlier fiscal years contributed a larger proportion of cumulative DDDs than the 2021–2022 period, indicating a declining utilization trajectory over time (Table 1).

## Discussion

In this 5-year dispensing-based analysis of cardiovascular outpatients at a national cardiac tertiary hospital in Tanzania, outpatient antibiotic consumption, quantified using the WHO ATC/DDD methodology, was low in absolute magnitude, yet consistently present across all fiscal years studied. Monthly utilization ranged from 0.014 to 0.049 DDD per 100 outpatients per day, for a cumulative total of 1.68 DDD per 100 outpatients per day across the full observation period. These values are substantially lower than the nationally reported bulk consumption estimates [6], underscoring the importance of patient-level dispensing data in interpreting antibiotic use. Outpatient antibiotic exposure in cardiology clinics may include prescriptions for non-infectious presentations, representing a potential target for optimizing unnecessary exposure.

International comparisons demonstrate that outpatient settings contribute materially to avoidable antibiotic use, particularly for syndromes in which bacterial infection is unlikely [24,25]. While the absolute quantities in this study were smaller, the relative distribution is meaningful: Access-group antibiotics dominated consumption, followed by minimal utilization of Watch-group antibiotics and Reserve-group antibiotics. This pattern aligns with stewardship expectations, but the presence of Watch-class outpatient exposure warrants attention. The minimal use of Reserve antibiotics is reassuring from a stewardship perspective and aligns with the WHO AWaRe targets that emphasize minimizing Watch and Reserve exposure in routine ambulatory care.

However, the Access proportion of 48.5% remained below the WHO Access-60 benchmark, indicating that outpatient prescribing optimization remains necessary. The increasing trend observed from late 2019 to early 2021 supports the need for continuous surveillance rather than single cross-sectional snapshots. In addition, the recurrent seasonal pattern observed in our time-series decomposition is consistent with seasonal variations in ambulatory antibiotic prescribing described in other outpatient datasets [24, 5].

Subgroup variation among patients reinforces this opportunity. Consumption was highest among individuals covered by the NHIF and among older adults, likely reflecting more frequent encounters, multimorbidity, and perceived clinical vulnerability. Private payers contributed a substantially smaller fraction of DDDs despite an appreciable attendance volume, consistent with the differential access to antibiotics previously described in East African mixed-financing models [14,17]. From a stewardship perspective, these patterns indicate that stewardship interventions should be tailored to the payer mix and patient age structure rather than assuming uniform utilization profiles. The narrow DU90 profile, dominated mainly by amoxicillin and azithromycin, is consistent with other Tanzanian national consumption reports based on defined daily dose per 1,000 inhabitants per day import data that similarly demonstrate that most outpatient antibiotic volume is concentrated in only a few molecules [6,7], as well as outpatient LMIC dispensing studies [26,28].

Comparable outpatient analyses from Ethiopia, Syria and Nigeria similarly show that ambulatory antibiotic exposure is common even in the absence of clear bacterial indications [26–29].

Importantly, pharmacist-led stewardship trials in Ethiopia demonstrated that up to half of ambulatory antibiotic prescriptions were avoidable [31], reinforcing the interpretation that small-volume outpatient exposure remains a meaningful target for stewardship. In addition, specialist hospital trend analyses have shown measurable correlations between consumption volumes and resistance trajectories over time [30], suggesting that even modest outpatient Watch-antibiotic exposure may have biologically relevant consequences. National time-series data from Japan and Korea have also documented significant shifts in outpatient antibiotic consumption at the population level [32,33]. Taken together, these studies reinforce the interpretation that the low absolute outpatient quantities observed at JKCI align with a broader pattern of ambulatory antibiotic use described in other settings.

Although the study has some limitations, these findings demonstrate measurable, though low-volume, antibiotic exposure among cardiovascular outpatients, supporting the formal integration of outpatient consumption monitoring into routine AMS dashboards. Linking dispensing to diagnosis codes and expanding this methodology to other Tanzanian hospitals would strengthen national stewardship capacity and enable better detection of subtle shifts in outpatient antibiotic utilization over time.

**Figure 1:**
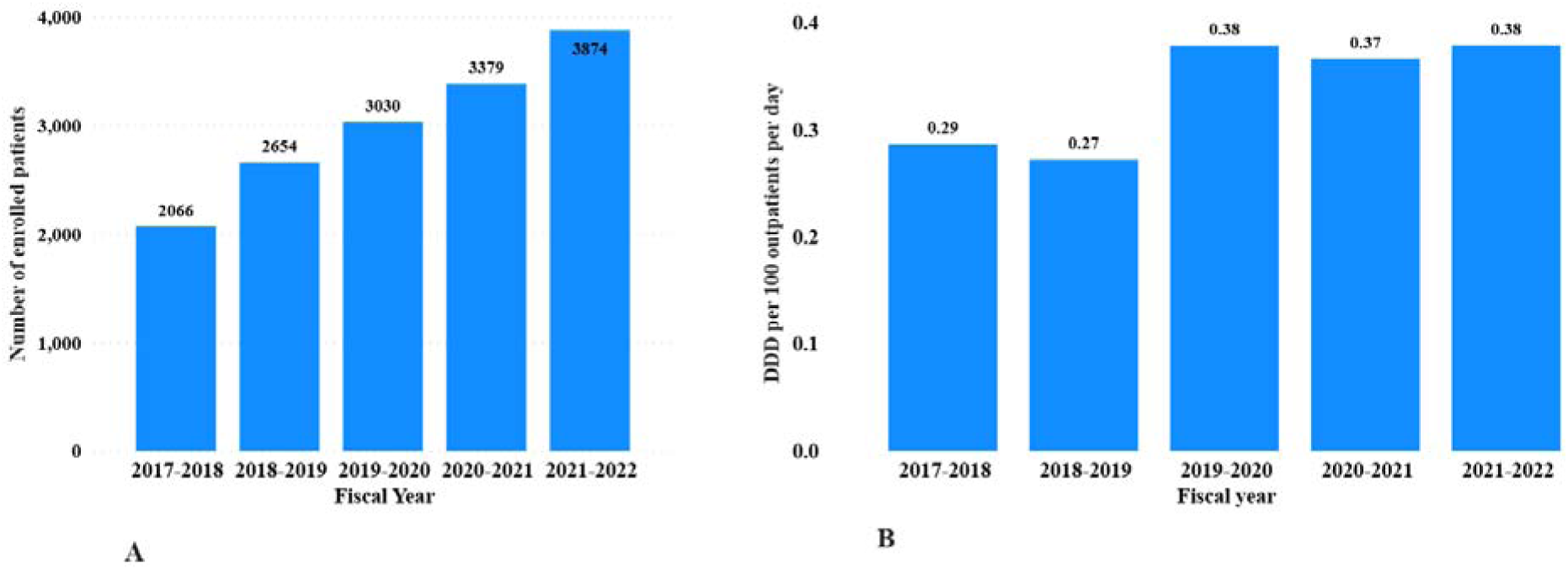
Annual outpatient attendance and antibiotic consumption at JKCI from fiscal years 2017–2018 to 2021–2022. Panel A: Number of cardiovascular outpatients dispensed at least one antibiotic prescription per fiscal year. Panel B: Annual antibiotic utilization expressed as DDD per 100 outpatients per day. Among ATC level 3 classes, beta-lactam penicillins (J01C) remained the dominant contributor to outpatient antibiotic utilization across all fiscal years, consistently recording the highest annual DDD per 100 outpatients per day, followed by other beta-lactams (J01D) and macrolides/lincosamides/streptogramins (J01F) (Figure 2).

**Figure 2:**
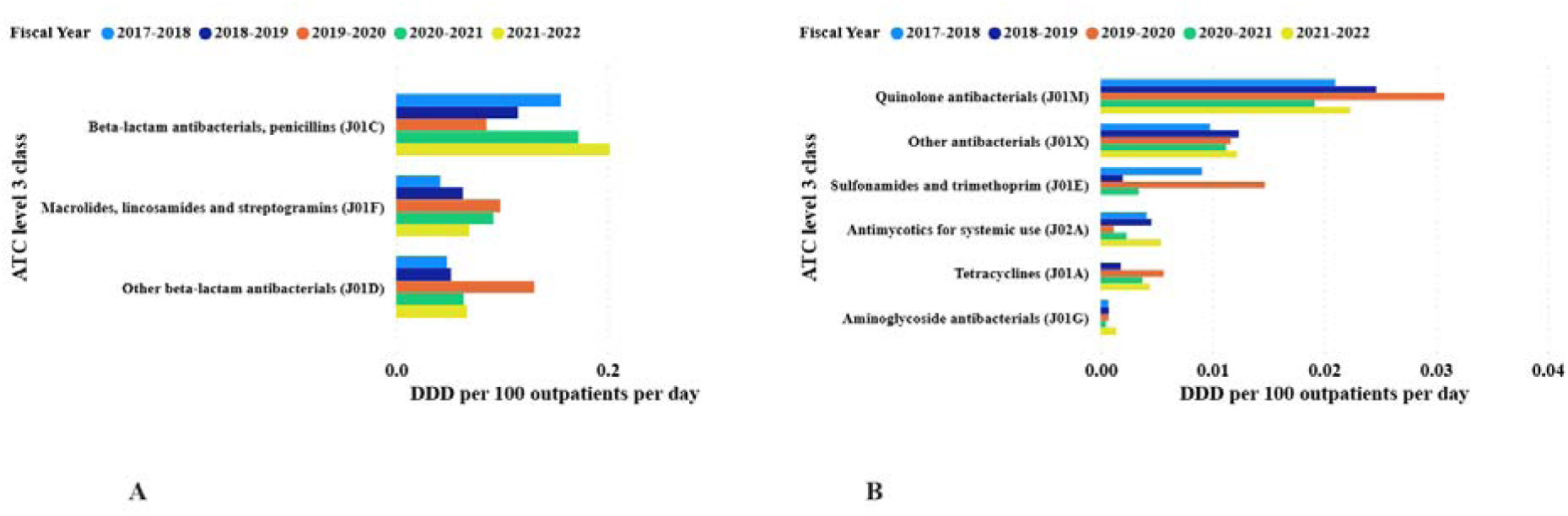
Antibiotic utilization by ATC level 3 classification across fiscal years. Panel A: Higher-volume ATC level 3 classes expressed as DDD per 100 outpatients per day. Panel B: Remaining lower-volume ATC level 3 classes. At ATC Level 4, third-generation cephalosporins (J01DD) displayed the highest DDD per 100 outpatients per day across fiscal years, followed by β-lactamase-resistant penicillins (J01CF) and macrolides (J01FA). In contrast, all remaining subclasses contributed comparatively smaller volumes (Figure 3).

**Figure 3:**
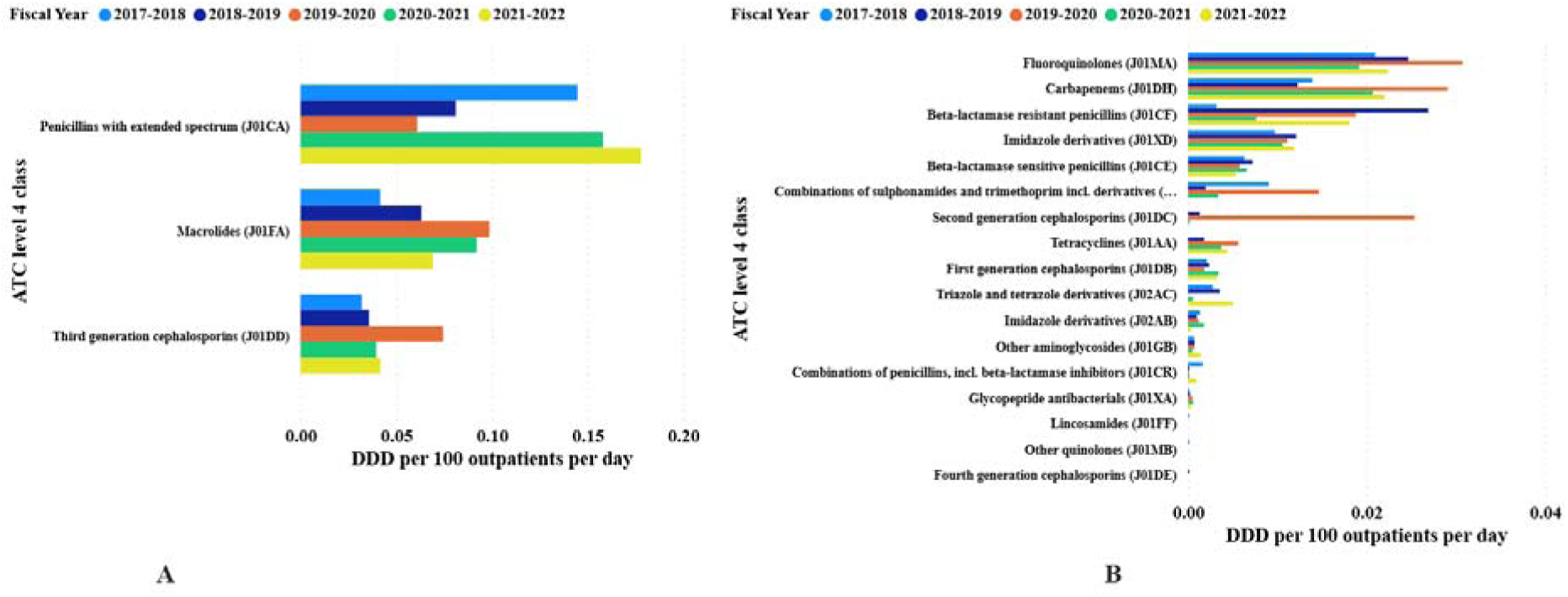
Antibiotic utilization by ATC Level 4 classification across fiscal years. Panel A: Higher-volume ATC level 4 subclasses expressed as DDD per 100 outpatients per day. Panel B: Remaining lower-volume ATC level 4 subclasses. At ATC level 5, outpatient antibiotic utilization was dominated by two antibiotics, amoxicillin (J01CA04) and azithromycin (J01FA10), which together accounted for the largest share of cumulative DDD per 100 outpatients per day across all fiscal years. The remaining 15 individual molecules contributed to comparatively small volumes (Figure 4). The DU90 segment comprised 11 molecules, accounting for 90.5% of cumulative consumption, and the DU50 threshold wa reached by the top three antibiotics (Supplementary Table 1).

**Figure 4:**
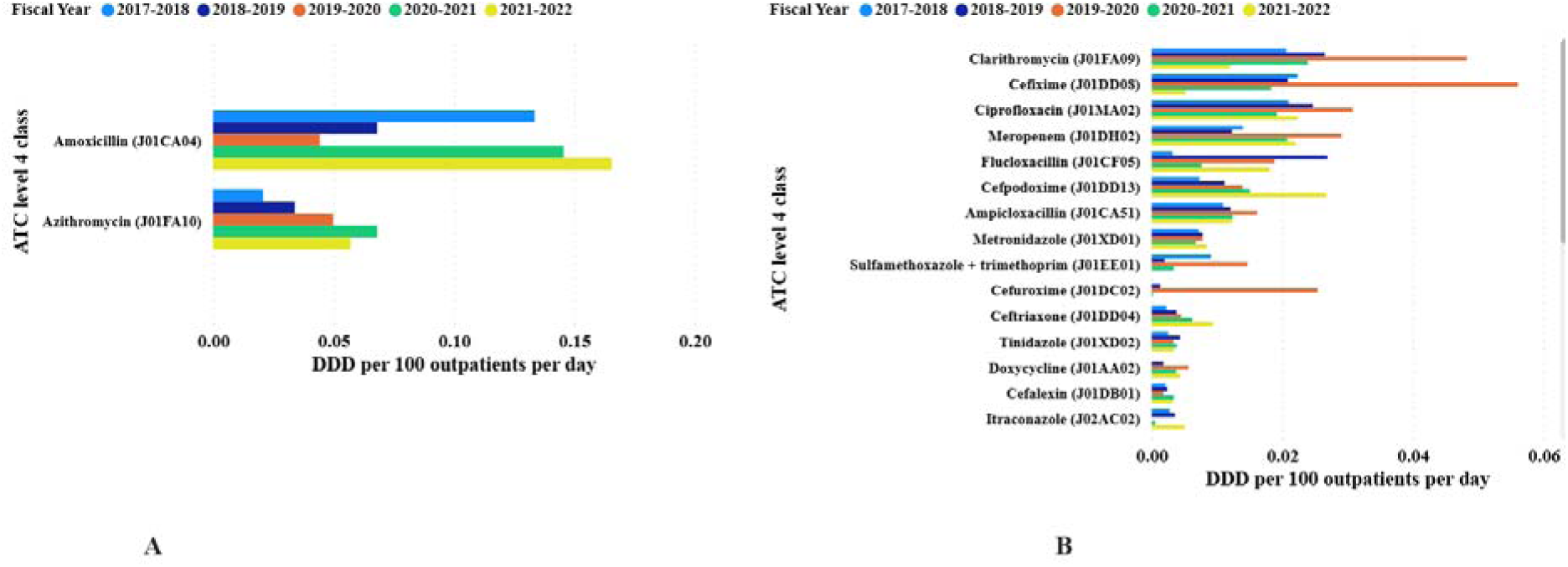
Antibiotic utilization by ATC level 5 classification across fiscal years. Panel A: Top two highest-volume level 5 molecules (amoxicillin and azithromycin) expressed a DDD per 100 outpatients per day. Panel B: Remaining lower-volume level 5 molecules (total n=17). Based on Option A values, Access antibiotics accounted for 0.816 DDD per 100 outpatients per day, Watch accounted for 0.765 DDD per 100 outpatients per day, and Reserve accounted for 0.100 DDD per 100 outpatients per day (Figure 5).

**Figure 5:**
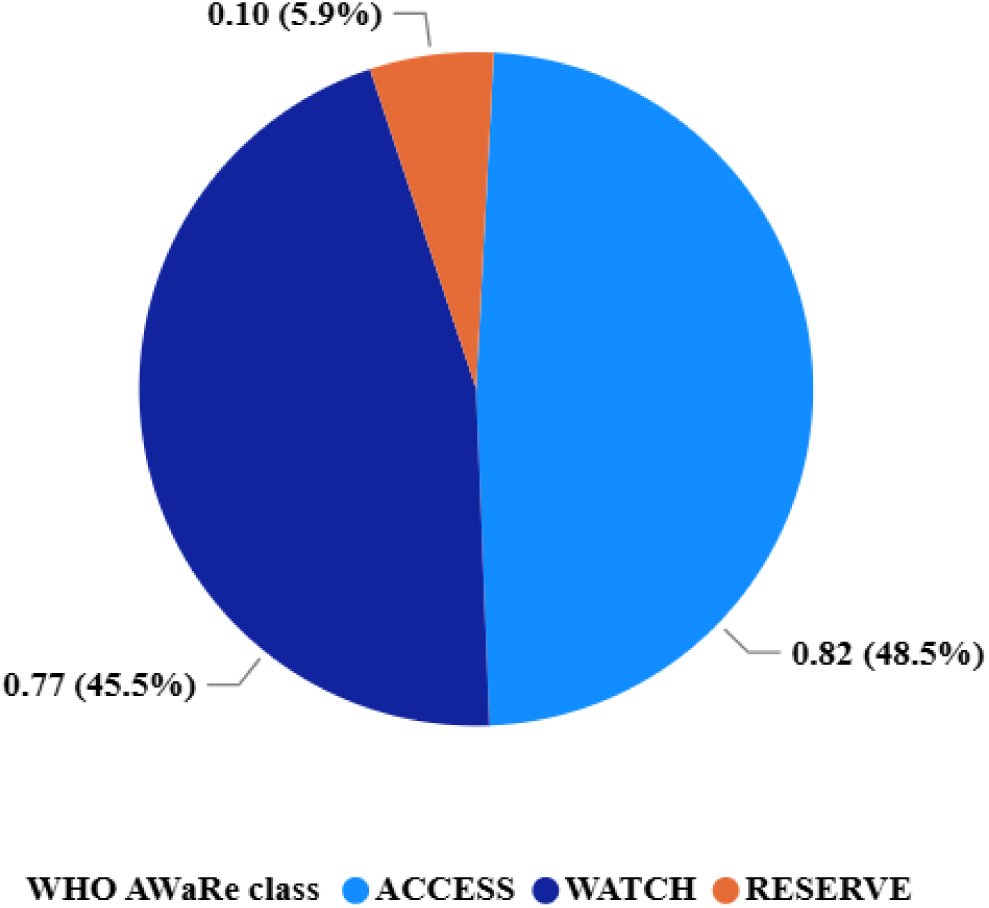
Outpatient antibiotic utilization by WHO AWaRe category across fiscal years. Monthly outpatient antibiotic consumption, expressed as DDD per 100 outpatients per day, fluctuated at low absolute levels over the five years, typically ranging from 0.014 to 0.049 DDD per 100 outpatients per day. Time-series decomposition revealed a gradual upward trend shift starting in late 2019, with peak monthly consumption in early 2021. A recurrent seasonal pattern was also detectable, with repeated short-cycle elevation peaks occurring at approximately similar calendar periods across successive years (Figure 6).

**Figure 6:**
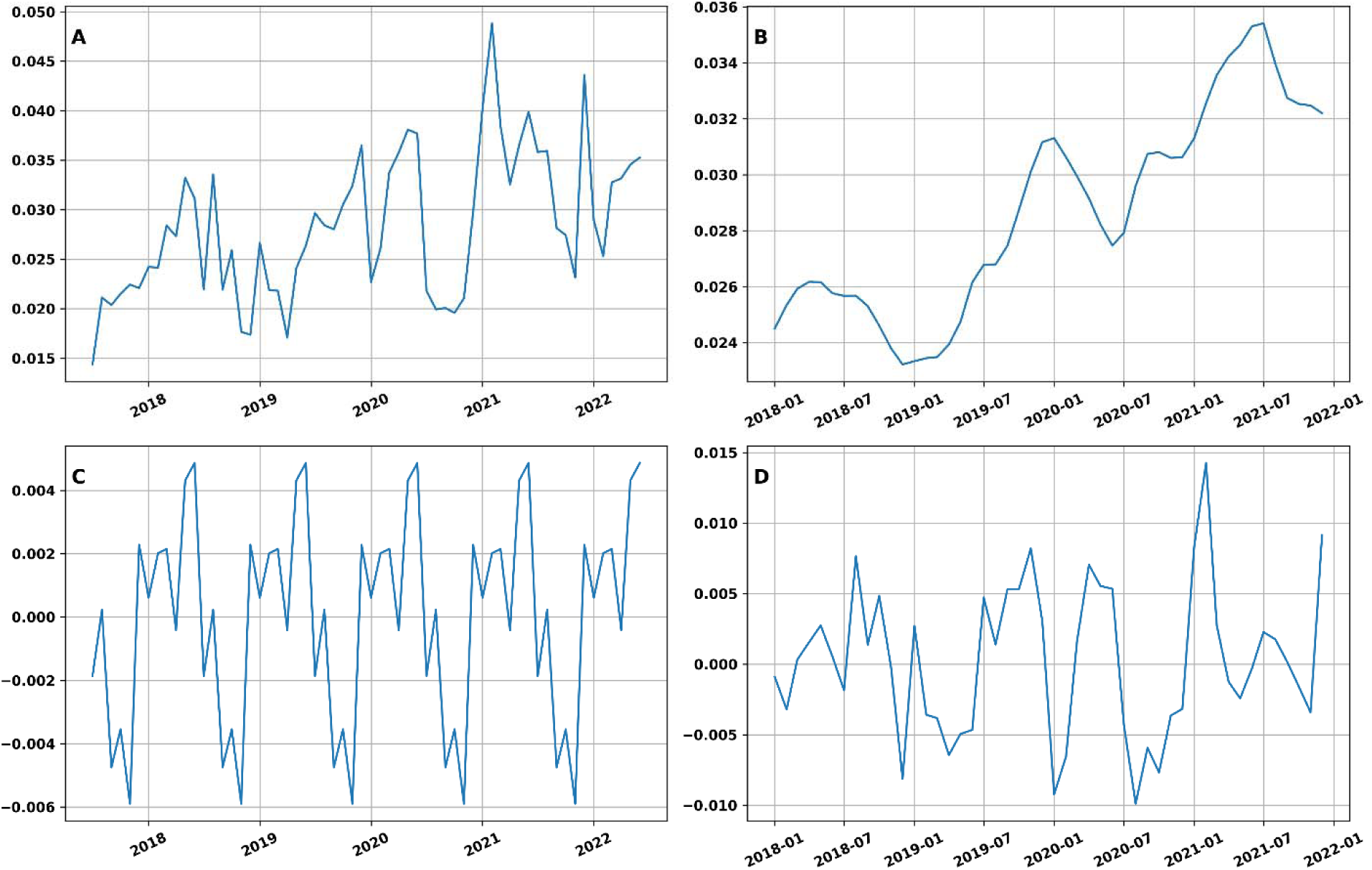
Time-series decomposition of monthly outpatient antibiotic consumption at the Jakaya Kikwete Cardiac Institute (JKCI), Dar es Salaam, Tanzania (July 2017–June 2022), expressed as DDD per 100 outpatients per day. Panels: **A** = Observed series; **B** = Trend; **C** = Seasonal component; **D** = Residuals The seasonal swing had an absolute amplitude of 0.0108 DDD per 100 outpatients per day, corresponding to approximately 38% relative variation between the peak and trough months.

## Conclusion

This study presents the first outpatient antibiotic utilization profile from a specialized cardiovascular referral center in Tanzania, utilizing the ATC/DDD methodology applied to real dispensing data. Although absolute outpatient exposure was low, antibiotics were consistently used across fiscal years, demonstrating a measurable seasonal pattern. Penicillin and beta-lactam/β-lactamase inhibitor combinations dominated prescriptions, accounting for nearly half of the cumulative exposure (45.5%). These proportional patterns indicate that stewardship attention is needed, even in non-infectious specialty outpatient clinics. Integrating routine antibiotic consumption monitoring into the existing stewardship dashboards at JKCI could provide an efficient mechanism to reinforce safer prescriptions, inform clinical audit cycles, and track shifts over time. Expanding similar dispensing-based surveillance to general outpatient care across Tanzania would establish national benchmarks for outpatient care and help strengthen implementation of the NAP AMR’s strategic objective to optimize antimicrobial use. These findings are consistent with those from other LMICs, where ambulatory antibiotic exposure, even at low absolute volumes, remains a relevant consideration for stewardship optimization.

## Limitations

This analysis used dispensing records, which reflect the actual medicines provided to patients; however, clinical indication data were unavailable, making it impossible to assess appropriateness. Consumption was quantified using DDDs, which are based on standard adult doses; therefore, values for children likely underestimated the actual exposure. Only systemic antibiotics were included and topical formulations were excluded. The study was conducted at a single center and in an outpatient-only setting; therefore, the generalizability of the findings beyond the tertiary cardiac referral setting in Dar es Salaam may be limited. Monthly fluctuations could reflect service patterns or diagnostic/insurance coding rather than true infection epidemiology. Finally, DDD metrics cannot be directly compared with DID values from population-level studies, and caution is required when interpreting cross-country comparisons.

## Recommendations

Routine outpatient antibiotic utilization reporting should be incorporated into the JKCI stewardship dashboards to enable continuous monitoring. Linking dispensing data with structured clinical indication fields (e.g., ICD-10 codes) would facilitate more targeted audits rather than exposure-only measurements. Similar outpatient surveillance should be expanded to additional tertiary and secondary hospitals to establish national reference values for comparison and validation. Within cardiology clinics, stewardship messaging should explicitly target AWaRe alignment, for example, limiting Watch antibiotics where acceptable Access alternatives exist. Finally, an annual time-series review of outpatient antibiotic data should continue to determine whether the early-2021 rise reappears and whether stewardship interventions alter the seasonal amplitude over time.

## Data Availability

All data produced in the present study are available upon reasonable request to the authors.

## Data Access Statement

Research data supporting this publication are available upon reasonable request from the corresponding author.

## Author Contributions

HAK, RZS and JAN contributed to study design, data acquisition, and data curation. RZS led data harmonization, ATC/DDD mapping oversight, and manuscript drafting.

JAN and JM performed data extraction and preliminary statistical analysis. ABM provided methodological oversight and manuscript review.

NM, RKM, and PRK reviewed the interpretation and contributed to contextualizing the findings. All authors reviewed and approved the final manuscript.

---

**Supplementary Table 1.** : Table of Defined Daily Dose (DDD) per 100 outpatients per day of antibiotics in the generic name (ATC level 5) classification of antibiotics showing consumption over time.

|  | **DDD per 100**<br>**outpatient/day/Fiscal Year** |  |  |  |  |  |  |  |
| --- | --- | --- | --- | --- | --- | --- | --- | --- |
| **Antibiotic generic**<br>**name(ATC level 5**<br>**code)** | **2017-2018** | **2018-**<br>**2019** | **2019-**<br>**2020** | **2020-**<br>**2021** | **2021-**<br>**2022** | **Molecule**<br>**total** | **Molecule**<br>**%** | **Cumulative**<br>**%** |
| Amoxicillin (J01CA04) | 0.133455 | 0.06807<br>7 | 0.04426<br>6 | 0.14551<br>6 | 0.16557<br>1 | 0.55688<br>5 | 32.8 | 32.8 |
| Azithromycin<br>(J01FA10) | 0.020762 | 0.03389<br>3 | 0.04986<br>2 | 0.06815 | 0.05711<br>5 | 0.22978<br>2 | 13.5 | 46.3 |
| Clarithromycin<br>(J01FA09) | 0.02058 | 0.02650<br>9 | 0.04823<br>2 | 0.02391<br>1 | 0.01201<br>7 | 0.13124<br>9 | 7.7 | 54.0 |
| Cefixime (J01DD08) | 0.022332 | 0.02084<br>1 | 0.05604<br>3 | 0.01829<br>5 | 0.00521<br>5 | 0.12272<br>6 | 7.2 | 61.3 |
| Ciprofloxacin<br>(J01MA02) | 0.020946 | 0.02466<br>5 | 0.03076<br>5 | 0.01916<br>2 | 0.02234<br>9 | 0.11788<br>7 | 6.9 | 68.2 |
| Meropenem (J01DH02) | 0.013947 | 0.01228<br>4 | 0.02909<br>1 | 0.02070<br>9 | 0.02201<br>1 | 0.09804<br>2 | 5.8 | 74.0 |
| Flucloxacillin<br>(J01CF05) | 0.003189 | 0.02691<br>6 | 0.01879 | 0.00763<br>9 | 0.01802<br>5 | 0.07455<br>9 | 4.4 | 78.4 |
| Cefpodoxime<br>(J01DD13) | 0.007305 | 0.01114<br>3 | 0.01390<br>8 | 0.01503 | 0.02675<br>7 | 0.07414<br>3 | 4.4 | 82.7 |
| Ampicloxacillin<br>(J01CA51) | 0.010928 | 0.01207<br>1 | 0.01615<br>3 | 0.01235<br>9 | 0.01229<br>8 | 0.06380<br>9 | 3.8 | 86.5 |
| Metronidazole<br>(J01XD01) | 0.007188 | 0.00777<br>5 | 0.00778<br>1 | 0.00676<br>8 | 0.00841<br>1 | 0.03792<br>3 | 2.2 | 88.7 |
| Phenoxymethylpenicilli<br>n-benzathine (J01CE10) | 0.006322 | 0.00678<br>5 | 0.00579<br>8 | 0.00657<br>5 | 0.00506<br>7 | 0.03054<br>7 | 1.8 | 90.5 |
| Sulfamethoxazole +<br>trimethoprim (J01EE01) | 0.009072 | 0.00198<br>1 | 0.01467<br>7 | 0.00339<br>6 |  | 0.02912<br>6 | 1.7 | 92.2 |
| Cefuroxime (J01DC02) |  | 0.00130<br>7 | 0.02538<br>5 | 0.00019<br>1 |  | 0.02688<br>3 | 1.6 | 93.8 |
| Ceftriaxone (J01DD04) | 0.002243 | 0.00379<br>7 | 0.00449<br>6 | 0.00620<br>3 | 0.00936<br>4 | 0.02610<br>3 | 1.5 | 95.3 |
| Tinidazole (J01XD02) | 0.00254 | 0.00435<br>2 | 0.00334<br>7 | 0.00384<br>5 | 0.00344<br>8 | 0.01753<br>2 | 1.0 | 96.4 |
| Doxycycline (J01AA02) |  | 0.00180<br>2 | 0.00563<br>2 | 0.00373<br>5 | 0.00438<br>4 | 0.01555<br>3 | 0.9 | 97.3 |
| Cefalexin (J01DB01) | 0.002105 | 0.00237<br>3 | 0.00182<br>2 | 0.00341<br>1 | 0.00322<br>3 | 0.01293<br>4 | 0.8 | 98.0 |
| Itraconazole (J02AC02) | 0.002755 | 0.00356<br>3 |  | 0.00053<br>4 | 0.00506<br>8 | 0.01192 | 0.7 | 98.7 |
| Miconazole (J02AB01) | 0.001347 | 0.00096<br>8 | 0.00118 | 0.00177<br>5 | 0.00032<br>6 | 0.00559<br>6 | 0.3 | 99.1 |
| Gentamicin (J01GB03) | 0.000692 | 0.00071<br>1 | 0.00069<br>5 | 0.00047<br>4 | 0.00139<br>6 | 0.00396<br>8 | 0.2 | 99.3 |
| Erythromycin<br>(J01FA01) | 0.00034 | 0.00267<br>9 | 0.00055<br>5 |  | 0.00004<br>6 | 0.00362 | 0.2 | 99.5 |
| Amoxicillin +<br>clavulanic (J01CR02) | 0.001658 | 0.00012<br>1 | 0.00008<br>6 | 0.00016<br>6 | 0.00092<br>2 | 0.00295<br>3 | 0.2 | 99.7 |
| Ampicillin (J01CA01) | 0.000293 | 0.00090<br>1 | 0.00051<br>8 | 0.00015<br>9 |  | 0.00187<br>1 | 0.1 | 99.8 |
| Vancomycin (J01XA01) | 0.000051 | 0.00022 | 0.00050 | 0.00059 | 0.00033 | 0.00170 | 0.1 | 99.9 |
|  |  |  | 1 | 4 | 6 | 2 |  |  |
| Phenoxymethylpenicillin (J01CE02) |  | 0.000456 |  |  | 0.000299 | 0.000755 | 0.0 | 100.0 |
| Ceftriaxone and beta-lactamase inhibitor (J01DD63) |  |  |  | 0.000057 | 0.00044 | 0.000497 | 0.0 | 100.0 |
| Cefoperazone + salbactam (J01DD62) | 0.000128 |  |  |  |  | 0.000128 | 0.0 | 100.0 |
| Clindamycin (J01FF01) | 0.000079 |  |  |  |  | 0.000079 | 0.0 | 100.0 |
| Nalidixic acid (J01MB02) | 0.000043 |  |  |  |  | 0.000043 | 0.0 | 100.0 |
| Amikacin (J01GB06) |  | 0.000014 | 0.00002 |  |  | 0.000034 | 0.0 | 100.0 |
| Cefepime (J01DE01) |  | 0.000031 |  |  |  | 0.000031 | 0.0 | 100.0 |
| Piperacillin + tazobactam (J01CR05) | 0.000001 | 0.000002 | 0.000001 | 0 | 0.000001 | 0.000005 | 0.0 | 100.0 |
| **Year Total** | **0.290301** | **0.276237** | **0.379604** | **0.368654** | **0.384089** | **1.698885** | **100.0** |  |

